# Immediate Block Is Not Stable Block: Early Mitral Isthmus Reconnection Despite Systematic Vein of Marshall Ethanol Infusion and Focal Pulsed Field Ablation With the Sphere-9™ Lattice-Tip Catheter

**DOI:** 10.64898/2026.08.28.26361686

**Authors:** Antoine Da Costa, Cédric Yvorel, Cécile Romeyer, Pierre Groussin, Alberto Barengo, Rayan Mohammed, Kasra Azarnoush, Nathalie Grand, Marouane Boukhris, Karim Benali

## Abstract

**Background:** Durable mitral isthmus (MI) block remains challenging in patients undergoing persistent atrial fibrillation (PeAF) ablation. Recent data using direct vein of Marshall (VoM) epicardial recordings have shown that endocardial pulsed field ablation (PFA) does not consistently achieve transmural MI lesions and that initially suppressed epicardial conduction may recover over time. Whether systematic VoM ethanol infusion (VoM-EI) combined with focal PFA overcomes this limitation and provides stable acute MI block remains unknown.

**Objectives:** To assess the incidence, timing, and procedural implications of early MI conduction recovery after systematic VoM-EI followed by focal PFA using the Sphere-9™ lattice-tip catheter.

**Methods:** This prospective, single-center observational study screened 55 consecutive patients undergoing a first catheter ablation for symptomatic PeAF with planned MI linear ablation. VoM-EI was systematically attempted before left atrial access and was successfully performed in 51 of 55 patients (92.7%), who constituted the study cohort. The lesion set comprised wide- antral pulmonary vein isolation, left atrial roof-line ablation, and MI-line ablation, all delivered using the Sphere-9™ lattice-tip catheter integrated with the Affera™ mapping and ablation system. After confirmation of bidirectional MI block, conduction was systematically reassessed following a standardized 30-minute waiting period. The primary endpoint was early MI conduction recovery.

**Results:** Mean age was 70.3 ± 8.2 years, and 36 patients (70.6%) were men. Initial bidirectional MI block was achieved in 50 of 51 patients (98.0%). During the 30-minute waiting period, MI conduction recovered in 9 of 50 patients with initial block (18.0%; 95% CI, 9.8%–30.8%), at a median of 16 minutes (IQR, 10–20; range, 8–23). Six of these 9 patients (66.7%) required additional coronary sinus (CS) ablation to restore block. Bidirectional MI block was re- established in all patients with conduction recovery, resulting in a final block rate of 50 of 51 patients (98.0%). Median procedure duration was 82 minutes (IQR, 73–95), and no major procedural complications occurred.

**Conclusions:** Immediate bidirectional MI block was not synonymous with stable block. Despite systematic VoM-EI followed by focal Sphere-9™ PFA, MI conduction recovered during a standardized 30-minute waiting period in approximately one in five patients, with two thirds requiring targeted CS ablation. Recovery occurred as late as 23 minutes, supporting reassessment beyond a 20-minute observation window. These findings extend recent evidence of incomplete MI transmurality after PFA by showing that even after systematic epicardial substrate modification with VoM-EI, apparently successful acute block may remain unstable. A standardized waiting period with systematic reassessment and targeted interrogation of residual CS conduction may therefore be warranted. Systematic invasive remapping will be required to determine whether waiting-period–guided re-ablation translates into durable chronic MI block.

Central Illustration:
Acute durability of mitral isthmus block after focal pulsed field ablation combined with first systematic vein of Marshall ethanol infusion. Systematic ethanol infusion followed by focal PFA with the Sphere-9™ lattice-tip catheter achieved acute bidirectional mitral isthmus block in 50 of 51 patients (98.0%). During a standardized 30-minute waiting period, conduction recovery occurred in 9 of 50 patients (18%), at a median of 16 minutes and as late as 23 minutes, and 6 of these 9 patients (66.7%) required targeted coronary sinus ablation. Final bidirectional block was achieved in 50 of 51 patients (98.0%), supporting systematic waiting-period reassessment after PFA-based mitral isthmus ablation.

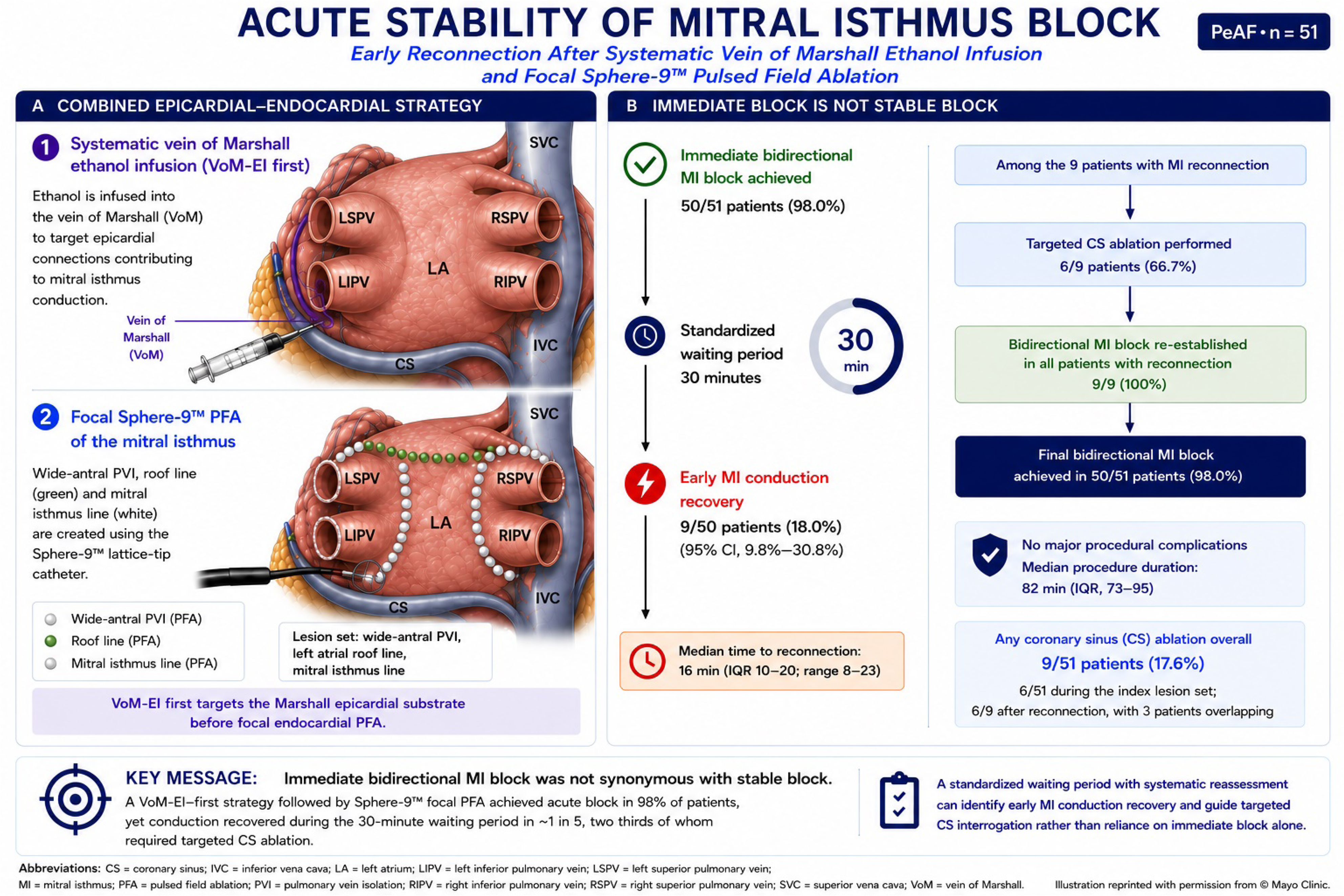

## INTRODUCTION

Catheter ablation is a cornerstone therapy for persistent atrial fibrillation (PeAF), yet the creation of durable left atrial linear lesions remains challenging (1–6). Mitral isthmus (MI) block is particularly difficult to achieve because of variable tissue thickness, complex fiber orientation, the cooling effect of the circumflex artery, coronary sinus (CS) muscular sleeves, and epicardial connections involving the vein and ligament of Marshall (1–6). Incomplete or non-durable MI block may promote recurrent perimitral macroreentrant tachycardia and repeat ablation procedures (2–6).

Vein of Marshall ethanol infusion (VoM-EI) facilitates MI block by targeting the Marshall bundle and associated epicardial connections while reducing the extent of endocardial ablation required (7–17). Zuo et al. showed that VoM-EI reduced MI conduction recovery after a standardized 30-minute waiting period from 35.1% to 6.7% (10), while invasive remapping studies have identified the mitral annulus and CS as frequent sites of late reconnection (11). These observations highlight that achievement of immediate bidirectional block does not necessarily establish lesion stability.

Pulsed field ablation (PFA) provides rapid, myocardial-selective lesion formation, but evidence regarding the transmurality and durability of PFA-created MI lines remains limited and heterogeneous (18–31). Patel et al. recently used direct epicardial VoM electrograms and VoM pacing to interrogate MI transmurality across three PFA platforms, including Sphere- 9™/Affera™. Endocardial PFA produced durable attenuation of VoM epicardial electrograms in only 52.1% overall and 67.7% with Affera, while VoM capture persisted after 20 minutes in 25.8% of Affera-treated patients (32). Pseudoblock due to persistent epicardial conduction was common, demonstrating that conventional endocardial criteria may overestimate true transmural MI block (32). These findings provide direct mechanistic evidence that apparently successful acute PFA effects at the MI may not uniformly reflect complete endocardial– epicardial interruption.

These observations raise a related but clinically distinct question. Patel et al. evaluated PFA before adjunctive VoM-EI, whereas a systematic VoM-EI–first strategy intentionally modifies the Marshall epicardial substrate before focal endocardial PFA. Whether apparently complete bidirectional MI block remains stable after this combined epicardial–endocardial approach is unknown. In particular, time-dependent conduction recovery or residual CS-related connections may become apparent only after an observation period despite initially successful block. A protocol-defined waiting period therefore provides a pragmatic assessment of acute block stability that is complementary to the direct evaluation of transmurality. To our knowledge, early MI conduction recovery has not been prospectively characterized after systematic first-line VoM-EI followed by focal Sphere-9™ PFA using protocol-mandated reassessment after documented bidirectional block.

We therefore prospectively evaluated the incidence, timing, and procedural implications of early MI conduction recovery after systematic first-line VoM-EI followed by focal PFA with the Sphere-9™ lattice-tip catheter, using protocol-defined reassessment after a standardized 30-minute waiting period. Secondary objectives were to assess acute efficacy, the need for targeted CS ablation, procedural efficiency, and safety.

## METHODS

### Study Design

This prospective, single-center observational study was conducted at the University Hospital of Saint-Étienne from October 2025 to August 2026 to evaluate the acute durability of MI block following focal PFA performed with the Sphere-9™ lattice-tip catheter and the Affera™ integrated mapping and ablation platform combined with systematic VoM-EI. Consecutive patients undergoing first-line catheter ablation for symptomatic PeAF, in whom MI linear ablation was planned as part of the ablation strategy, were prospectively enrolled. Clinical, imaging, electrophysiological, procedural, and follow-up data were prospectively recorded in a dedicated institutional database. The study complied with the Declaration of Helsinki and was approved by the local Institutional Review Board. All patients provided written informed consent for catheter ablation and for the anonymous use of their clinical data for research purposes. The study is reported in accordance with the STROBE statement for observational studies.

### Study Population and Analysis Set

Patients with symptomatic PeAF refractory to antiarrhythmic drug therapy were screened for eligibility. PeAF was defined as continuously sustained AF lasting more than 7 days, including episodes terminated by pharmacological or electrical cardioversion after 7 days. Patients were eligible if they were 18 years of age or older, had symptomatic PeAF refractory to at least one antiarrhythmic drug, and were undergoing their first ablation procedure for PeAF. Exclusion criteria were paroxysmal AF; long-standing persistent AF lasting more than 3 years; age below 18 years; intracardiac thrombus diagnosed by preprocedural transesophageal echocardiography and/or cardiac computed tomography; lack of written informed consent; and contraindication to ethanol infusion.

Fifty-five consecutive patients were enrolled. Because the question addressed here concerns the stability of MI block after the Marshall epicardial substrate has actually been treated, the analysis set was restricted a priori to patients in whom VoM-EI was completed. The vein of Marshall was identified at venography and successfully alcoholized in 51 of the 55 enrolled patients (92.7%); the remaining 4 patients (7.3%), in whom the vein could not be identified, were excluded from the analysis. All results reported below therefore refer to these 51 patients, and the 92.7% figure is retained as the feasibility of the systematic VoM-EI strategy in the screened population.

### Preprocedural Evaluation

All patients were required to continue oral anticoagulation, including direct oral anticoagulants or vitamin K antagonists, without interruption for at least 3 weeks before the procedure and until the day of the procedure. Before the procedure, all patients underwent comprehensive clinical evaluation including medical history, physical examination, 12-lead electrocardiography, transthoracic echocardiography, and transesophageal echocardiography or cardiac computed tomography according to institutional practice. Baseline demographic characteristics, cardiovascular risk factors, echocardiographic parameters, and laboratory findings were prospectively collected.

### Ablation System and Pulsed Field Settings

Mapping and ablation were performed with the Affera™ mapping and ablation system using the Sphere-9™ catheter (Medtronic, Minneapolis, MN, USA). This 7.5F dual-energy catheter carries a compressible 9-mm lattice-shaped nitinol tip incorporating surface mini-electrodes and thermocouples, allowing ultra-high-density mapping and focal point-by-point delivery of either temperature-controlled irrigated radiofrequency or a proprietary biphasic pulsed field waveform from the same wide-footprint electrode, each application lasting a few seconds (21, 22). In the present study, all endocardial lesions were created with pulsed field energy only; the radiofrequency mode was not used.

### Electrophysiological Procedure

All procedures were performed under general anesthesia. Following ultrasound-guided femoral venous access, a decapolar catheter was positioned within the coronary sinus. A single transseptal puncture was performed under fluoroscopic guidance. Intravenous unfractionated heparin was administered immediately after transseptal access to maintain an activated clotting time greater than 300 seconds throughout the procedure. Electroanatomical mapping was performed using the Affera™ high-density mapping platform. In patients not in sinus rhythm at the beginning of the procedure, electrical cardioversion was systematically performed before mapping. Three-dimensional left atrial geometry and baseline activation and voltage maps were acquired during coronary sinus pacing at a cycle length of 600 ms, allowing precise delineation of the pulmonary veins, left atrial appendage, mitral annulus, posterior wall, and coronary sinus. The pulmonary vein isolation–linear plan workflow was implemented in the following sequence: VoM-EI first, performed before left atrial access; then transseptal puncture; left pulmonary vein isolation; MI ablation until bidirectional block was achieved; right pulmonary vein isolation; and finally roof-line ablation until block was confirmed. The MI line was reassessed during and after a 30-minute waiting period. The ablation strategy comprised, in all patients, wide-antral pulmonary vein isolation in addition to the mitral isthmus line and a left atrial roof line. All endocardial lesions were delivered with the Sphere-9™ lattice-tip catheter in a point-by-point fashion. Pulmonary vein isolation was confirmed by entrance and exit block, and roof-line block was confirmed by differential pacing and activation mapping.

### Vein of Marshall Ethanol Infusion

Prior to left atrial access, ethanol infusion into the VoM was performed as previously described, with minor modifications. The coronary sinus was cannulated with a large deflectable sheath (Agilis™, Abbott, St. Paul, MN, USA). Coronary sinus opacification was performed with a 7.5F thermodilution balloon catheter (Swan-Ganz, Edwards Lifesciences, Irvine, CA, USA) in an anteroposterior view, in order to identify the presence or absence of the VoM. A left internal mammary artery guide catheter was then used for angiographic contrast injection and catheterization of the VoM ostium, which was subselectively cannulated. An angioplasty guidewire (SION blue®, Asahi Intecc, Santa Ana, CA, USA) was advanced into the VoM, and a preloaded over-the-wire angioplasty balloon 1.5, 2, or 2.5 mm in diameter (Sprinter™ OTW, Medtronic) was advanced into the proximal VoM and inflated. VoM anatomy and occlusion were confirmed by venography. Absolute (100%) ethanol was then slowly injected through the central balloon lumen at a rate of 3 mL per minute initially and at a rate of 2ml for the last 4 ml, after which the balloon was deflated. The injected volume was 10 mL whenever the vein accommodated it.

### Mitral Isthmus Ablation

After completion of VoM-EI, MI ablation was performed using the Sphere-9™ focal PFA catheter. Point-by-point applications were delivered to create a continuous linear lesion extending from the lateral mitral annulus to the left inferior pulmonary vein. Ablation continued until bidirectional block was achieved. Clockwise MI block was confirmed if pacing from the left atrial appendage produced a proximal-to-distal activation sequence on the decapolar catheter positioned in the coronary sinus, and if differential pacing from the left atrial anterior wall resulted in progressive shortening of conduction time to the proximal CS. Counter-clockwise block was assessed by pacing from the 1–2 and 3–4 electrode pairs of the CS catheter and was confirmed by a distal-to-proximal activation sequence along the lateral left atrium with a corresponding prolongation of the transisthmus conduction time, corroborated by activation mapping whenever the pacing criteria were equivocal. Whenever residual conduction persisted after completion of the endocardial lesion set, it was mapped by activation and differential pacing to confirm epicardial breakthrough, and additional focal PFA applications were delivered. Complementary pulsed field applications within the CS and/or along the endocardial aspect of the mitral isthmus were performed at the operator’s discretion whenever required to achieve complete bidirectional conduction block. When applications were delivered from within the CS, the lattice electrode was positioned opposite the endocardial line under combined fluoroscopic and electroanatomical guidance. Only pulsed field energy was delivered within the CS; each application consisted of 1 second of unipolar biphasic pulsed field delivery, and 2 to 4 applications were required. No systematic intravenous nitroglycerin dose was administered before coronary sinus applications, but continuous 12-lead ST-segment monitoring was performed throughout the procedure and patients underwent continuous ECG monitoring until discharge on the following day.

### Standardized Waiting Period

Following confirmation of complete bidirectional MI block, a standardized 30-minute waiting period was systematically performed in all patients in whom acute block had been obtained. During and at the end of this period, pacing maneuvers and, whenever necessary, activation mapping was repeated to reassess conduction across the MI. Acute recovery of conduction was defined as any restoration of electrical conduction across the MI after initial confirmation of complete bidirectional block. Patients in whom bidirectional block was never obtained could not enter the waiting period and were therefore not counted as reconnection events. When reconnection was identified, additional focal PFA applications were delivered — endocardially and, when required, within the CS — until bidirectional block was re-established.

### Study Endpoints

The primary endpoint was acute lesion durability, defined as the absence of MI conduction recovery after the standardized 30-minute waiting period, assessed among patients in whom acute bidirectional block had been achieved. Secondary endpoints included the rate of acute bidirectional MI block, the requirement for complementary endocardial or coronary sinus ablation, the time to conduction recovery, procedure duration, fluoroscopy time, radiation exposure, and major procedural complications. Major complications were defined as cardiac tamponade, stroke or transient ischemic attack, coronary spasm, atrio-esophageal fistula, need for emergency cardiac surgery, vascular complication requiring intervention, and procedure-related death.

### Statistical Analysis

Continuous variables were tested for normality using the Shapiro–Wilk test. Normally distributed variables are presented as mean ± standard deviation, whereas non-normally distributed variables are expressed as median and interquartile range (IQR). Categorical variables are reported as counts and percentages. Confidence intervals for proportions were calculated using the Wilson score method. Because of the prospective single-arm design, analyses were primarily descriptive. Exploratory comparisons between patients with and without acute conduction recovery were performed using the Mann–Whitney U test for continuous variables and the Fisher exact test for categorical variables. Given the small number of observed events, no multivariable model was fitted, and these comparisons should be regarded as hypothesis-generating. Missing data were not imputed and all analyses were performed on an available-case basis; denominators are reported throughout. Statistical significance was defined as a two-sided P value < 0.05.

## RESULTS

### Study Population

Fifty-five consecutive patients with symptomatic PeAF undergoing first-line catheter ablation were enrolled. The vein of Marshall was identified and successfully alcoholized in 51 patients (92.7%), who constitute the analysis set; the 4 patients (7.3%) in whom the vein could not be identified at venography were excluded (Figure 1). Baseline clinical characteristics of the 51 analysed patients are summarized in Table 1. Mean age was 70.3 ± 8.2 years and 36 patients (70.6%) were male. Hypertension was present in 31 patients (60.8%), diabetes mellitus in 7 (13.7%), dyslipidemia in 20 (39.2%), coronary artery disease in 13 (25.5%), previous stroke or transient ischemic attack in 7 (13.7%), and heart failure in 37 (72.5%). The median CHA₂DS₂- VA score was 3 (IQR 2–4). Median AF duration before ablation was 12 months (IQR 12–24). Mean left ventricular ejection fraction was 54.4 ± 13.0% and median indexed left atrial volume was 55 mL/m² (IQR 44–76). Moderate or greater mitral regurgitation was present in 7 patients (13.7%), and amiodarone was used in 24 patients (47.1%) before the procedure.

**Figure 1.**
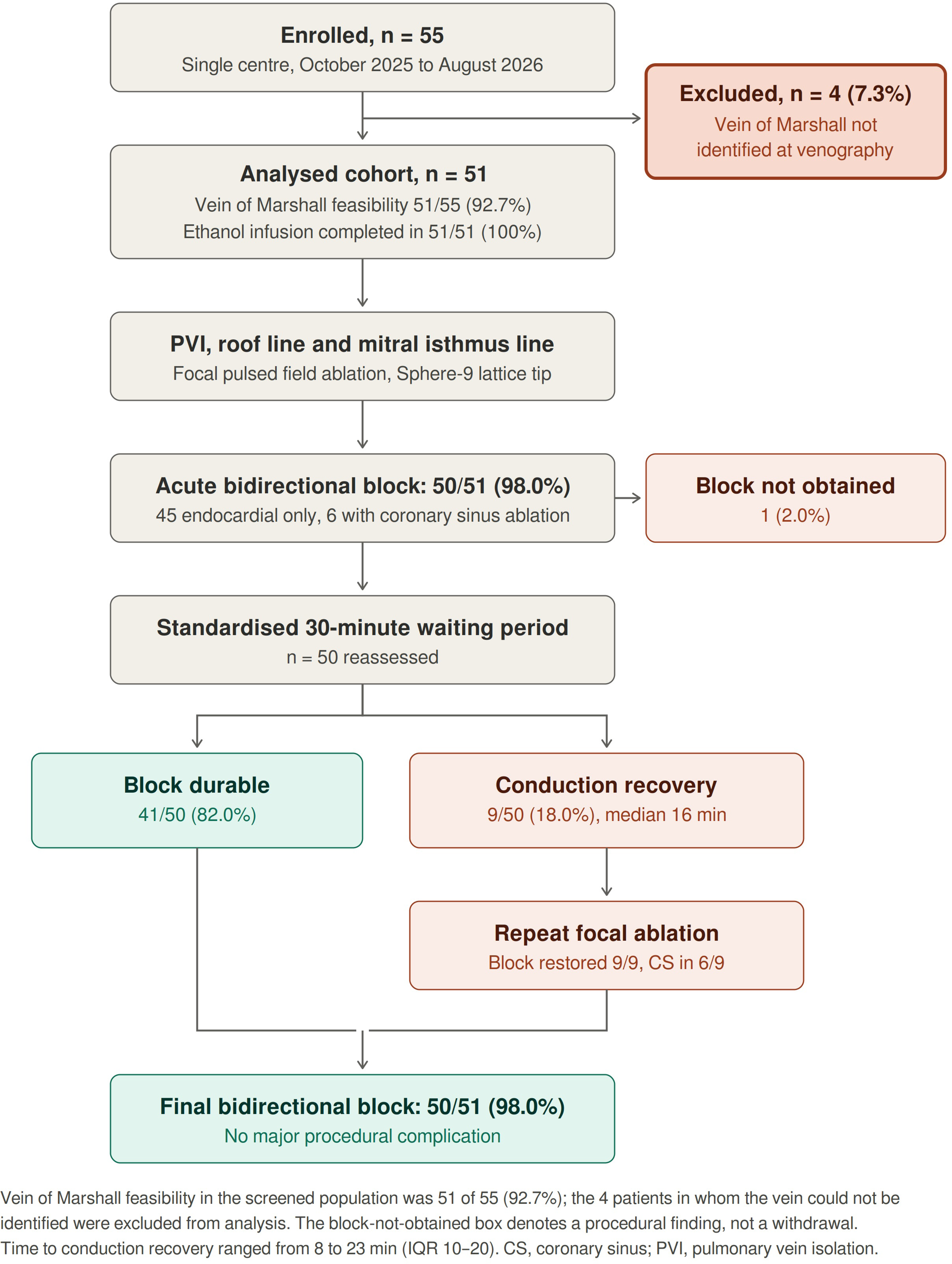
Study flow chart. Enrollment of 55 consecutive patients, exclusion of the 4 patients (7.3%) in whom the vein of Marshall could not be identified at venography, acute bidirectional block, waiting-period reassessment, and final outcome in the 51 analyzed patients. Vein of Marshall feasibility in the screened population was 51 of 55 (92.7%). Time to conduction recovery ranged from 8 to 23 minutes (IQR 10–20). CS, coronary sinus; PVI, pulmonary vein isolation.

**Table 1.** Baseline Characteristics.

| Variable | Analysed cohort (n = 51) |
| --- | --- |
| Age, years | 70.3 ± 8.2 |
| Male sex | 36 (70.6%) |
| Body mass index, kg/m<sup>2</sup> (n = 50) | 29.4 ± 6.9 |
| Hypertension | 31 (60.8%) |
| Diabetes mellitus | 7 (13.7%) |
| Current or former smoking | 12 (23.5%) |
| Dyslipidemia | 20 (39.2%) |
| Heart failure | 37 (72.5%) |
| Prior stroke or transient ischemic attack | 7 (13.7%) |
| Vascular disease | 17 (33.3%) |
| Coronary artery disease | 13 (25.5%) |
| CHA<sub>2</sub>DS<sub>2</sub>-VA score | 3 (2–4) |
| Persistent atrial fibrillation | 51 (100.0%) |
| Atrial fibrillation duration, months | 12 (12–24) |
| First-line (de novo) ablation procedure | 50 (98.0%) |
| Left ventricular ejection fraction, % | 54.4 ± 13.0 |
| Left atrial surface area, cm<sup>2</sup> | 27.0 ± 4.2 |
| Indexed left atrial volume, mL/m<sup>2</sup> | 55 (44–76) |
| Moderate or greater mitral regurgitation | 7 (13.7%) |
| Obstructive sleep apnea | 18 (35.3%) |
| Valve prosthesis | 4 (7.8%) |
| Beta-blocker therapy | 43 (84.3%) |
| Amiodarone therapy | 24 (47.1%) |
| Class Ic antiarrhythmic therapy | 5 (9.8%) |

Values are mean ± SD, median (interquartile range), or n (%), as appropriate. Percentages use available-case denominators; body mass index was available in 50 patients. CHA<sub>2</sub>DS<sub>2</sub>-VA = congestive heart failure, hypertension, age ≥ 75 years, diabetes mellitus, prior stroke or transient ischemic attack, vascular disease, and age 65–74 years (2024 ESC formulation, without the sex category).

### Procedural Characteristics

Procedural data are summarized in Table 2. Median skin-to-skin procedure duration was 82 minutes (IQR 73–95). Median total fluoroscopy time was 9.1 minutes (IQR 6.3–12.4), and median radiation exposure was 130.0 mGy (IQR 65.3–282.0). Median total PFA delivery duration was 29 minutes (IQR 21–41), corresponding to a median of 83 applications (IQR 62– 103) per procedure, including 16 (IQR 10–20) along the MI and 10 (IQR 8–12) along the roof line. Mean MI length was 3.2 ± 0.9 cm. VoM-EI was successfully completed in all 51 patients, with a median ethanol volume of 9 mL (IQR 6–10; range 5–10), a median VoM-EI procedural duration of 12 minutes (IQR 10–15), and a median VoM-EI fluoroscopy time of 2.3 minutes (IQR 1.4–4.1). Complementary PFA within the CS was required in 6 patients (11.8%) during the initial lesion set. Among the 6 patients requiring additional CS ablation after waiting-period reconnection, 3 had already undergone CS ablation during the initial lesion set. Overall, 9 distinct patients (17.6%) underwent CS ablation during the procedure.

**Table 2.**
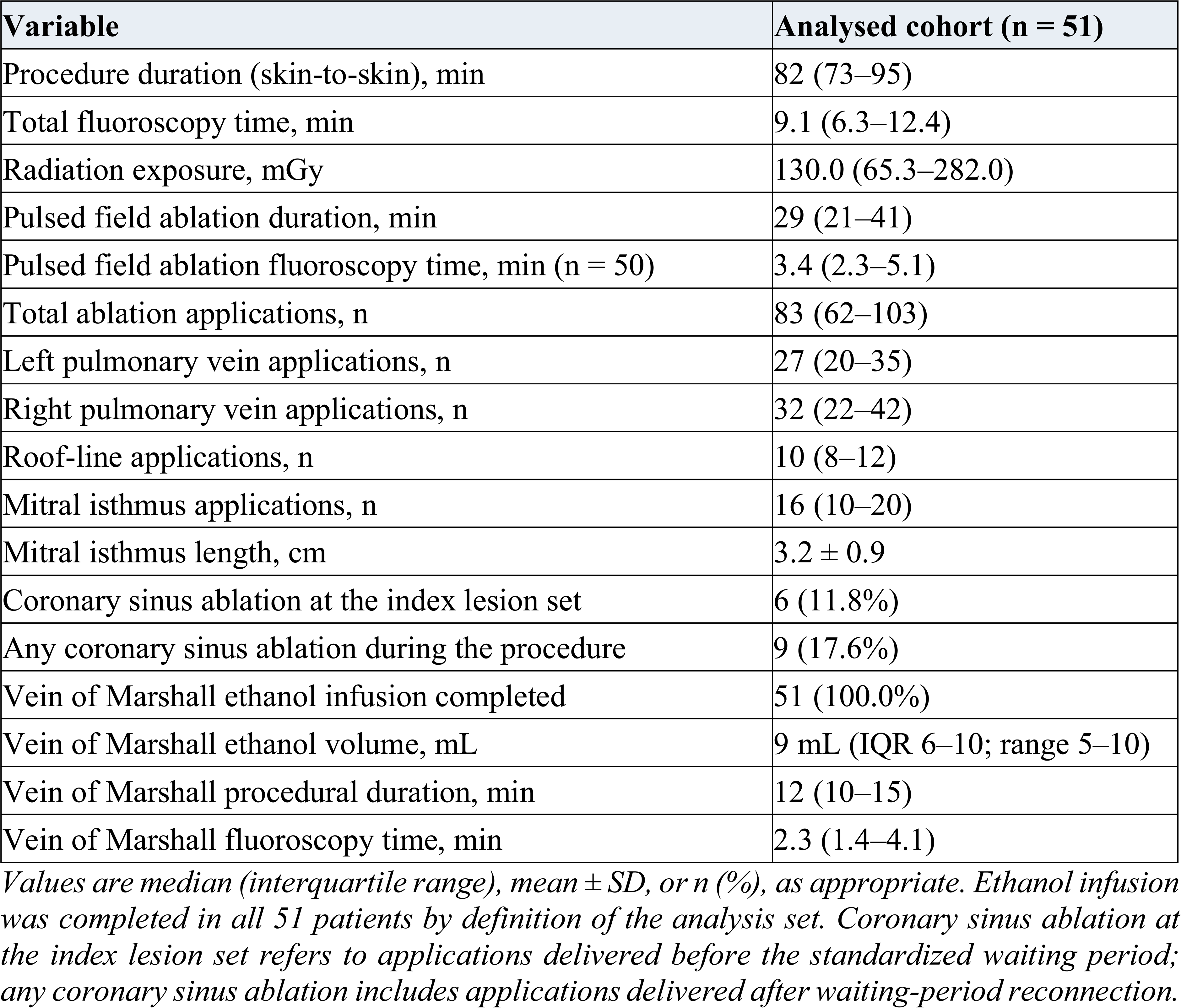
Procedural Characteristics.

| Variable | Analysed cohort (n = 51) |
| --- | --- |
| Procedure duration (skin-to-skin), min | 82 (73–95) |
| Total fluoroscopy time, min | 9.1 (6.3–12.4) |
| Radiation exposure, mGy | 130.0 (65.3–282.0) |
| Pulsed field ablation duration, min | 29 (21–41) |
| Pulsed field ablation fluoroscopy time, min (n = 50) | 3.4 (2.3–5.1) |
| Total ablation applications, n | 83 (62–103) |
| Left pulmonary vein applications, n | 27 (20–35) |
| Right pulmonary vein applications, n | 32 (22–42) |
| Roof-line applications, n | 10 (8–12) |
| Mitral isthmus applications, n | 16 (10–20) |
| Mitral isthmus length, cm | 3.2 ± 0.9 |
| Coronary sinus ablation at the index lesion set | 6 (11.8%) |
| Any coronary sinus ablation during the procedure | 9 (17.6%) |
| Vein of Marshall ethanol infusion completed | 51 (100.0%) |
| Vein of Marshall ethanol volume, mL | 9 mL (IQR 6–10; range 5–10) |
| Vein of Marshall procedural duration, min | 12 (10–15) |
| Vein of Marshall fluoroscopy time, min | 2.3 (1.4–4.1) |

Values are median (interquartile range), mean ± SD, or n (%), as appropriate. Ethanol infusion was completed in all 51 patients by definition of the analysis set. Coronary sinus ablation at the index lesion set refers to applications delivered before the standardized waiting period; any coronary sinus ablation includes applications delivered after waiting-period reconnection.

### Acute Procedural Efficacy

Acute procedural outcomes are summarized in Table 3. Acute bidirectional MI block was achieved in 50 of 51 patients (98.0%) and was obtained with endocardial ablation alone in 45 of 51 patients (88.2%); complementary applications within the CS were required at the index lesion set in 6 patients (11.8%). Complete roof-line block was obtained in all patients (100%). One patient exhibited persistent residual mitral isthmus conduction despite additional PFA including within the coronary sinus; because bidirectional block was never obtained in this patient, he could not enter the waiting period. All patients were in sinus rhythm at the completion of the procedure.

**Table 3.** Acute Procedural Outcomes.

| Variable | Analysed cohort (n = 51) |
| --- | --- |
| Acute bidirectional mitral isthmus block | 50/51 (98.0%) |
| Block achieved with endocardial ablation alone | 45/51 (88.2%) |
| Coronary sinus ablation required at the index lesion set | 6/51 (11.8%) |
| Roof-line block achieved | 51/51 (100.0%) |
| Sinus rhythm at procedure completion | 51/51 (100.0%) |
| Any major procedural complication | 0/51 (0.0%) |

Values are n/N (%). Endocardial-only block (n = 45), coronary sinus–assisted block (n = 6) and persistent conduction (n = 1) account for all 51 patients.

### Acute Durability of Mitral Isthmus Block

The 50 patients in whom acute bidirectional block was obtained underwent the standardized 30-minute waiting period. On repeated electrophysiological assessment during and at the end of the waiting period, acute recovery of MI conduction occurred in 9 of 50 patients (18%; 95% CI, 9.8–30.8), corresponding to a primary endpoint of acute lesion durability in 41 of 50 patients (82.0%). The median time from initial demonstration of block to documented conduction recovery was 16 minutes (IQR, 10–20), with a full range of 8 to 23 minutes; two patients reconnected beyond 20 minutes. Whenever reconnection was identified, additional focal PFA applications were delivered until complete bidirectional block was re-established, which was achieved in all 9 patients (100%). Complementary CS ablation was required to restore block in 6 of these 9 patients (66.7%). Consequently, bidirectional mitral isthmus block at the end of the procedure was present in 50 of 51 patients (98.0%). Acute durability data are detailed in Table 4.

**Table 4.** Acute Lesion Durability After the Standardized 30-Minute Waiting Period.

| Variable | Value |
| --- | --- |
| Patients entering the waiting-period assessment | 50 |
| Durable block after the waiting period (primary endpoint) | 41/50 (82.0%) |
| Mitral isthmus conduction recovery | 9/50 (18.0%; 95% CI 9.8–30.8) |
| Time from initial block to conduction recovery, min | 16 (10–20) |
| Full range of time to conduction recovery, min | 8–23 |
| Reconnection cases requiring coronary sinus ablation<br>of whom already treated in the CS at the index lesion set | 6/9 (66.7%)<br>3 |
| Block restored after additional ablation | 9/9 (100.0%) |
| Final complete mitral isthmus block | 50/51 (98.0%) |

Values are n/N (%) or median (interquartile range), as appropriate. Durability was evaluated among the 50 patients in whom acute bidirectional block was obtained; the single patient in whom block was never achieved could not enter the waiting period. Confidence intervals use the Wilson score method. The dataset did not contain a populated field distinguishing endocardial from epicardial reconnection; no such classification was imputed.

**Table 5.**
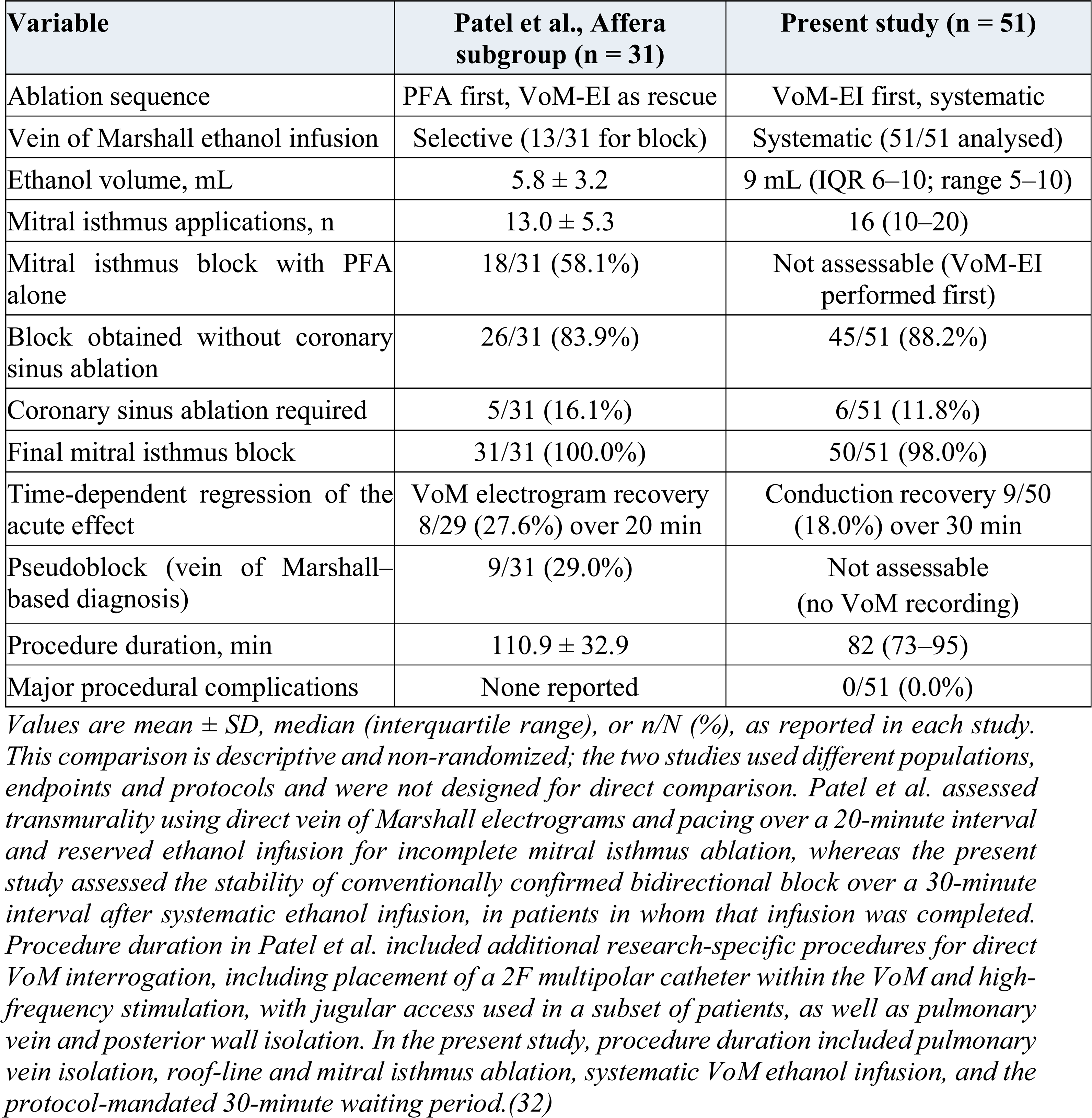
Descriptive Comparison With the Affera Subgroup of Patel et al.

### Predictors of Acute Mitral Isthmus Reconnection

In an exploratory comparison, patients with reconnection had a larger left atrium on echocardiography (indexed left atrial volume 76 (61–100) vs 52 (42–68) mL/m², P = 0.011; left atrial area 29 (28–32) vs 25 (24–29) cm², P = 0.014). Coronary sinus ablation at the index lesion set was numerically more frequent in patients who subsequently reconnected (3/9 (33.3%) vs 3/41 (7.3%), P = 0.063), a finding most plausibly reflecting a more challenging isthmus rather than an effect of CS ablation itself. Mitral isthmus length, total and endocardial application counts, electroporation time, left ventricular ejection fraction, ethanol volume and age did not differ between groups (all P > 0.33). Given only 9 events and multiple comparisons, these analyses are hypothesis-generating and no adjustment for multiplicity was applied.

### Procedural Safety

No major procedural complication occurred. Specifically, there were no cases of cardiac tamponade, stroke or transient ischemic attack, coronary artery spasm, atrio-esophageal fistula, need for emergency cardiac surgery, or procedure-related death. No complication related to VoM-EI was observed, and no vascular complication required intervention.

## DISCUSSION

### Main Findings

This prospective study specifically evaluated early conduction recovery after confirmed bidirectional MI block using a systematic first-line VoM-EI strategy followed by focal Sphere- 9™ PFA and protocol-defined 30-minute reassessment. The principal finding is that acute block and stable block are not synonymous. Although initial bidirectional MI block was achieved in 50 of 51 patients (98.0%), conduction recovered in 9 of 50 (18.0%) during the waiting period, at a median of 16 minutes and as late as 23 minutes. Block was restored in all 9 patients, but 6 (66.7%) required additional CS ablation. Thus, a high immediate success rate can conceal early conduction instability even after a deliberately combined epicardial– endocardial strategy and systematic treatment of the Marshall substrate.

A second observation concerns the timing of reassessment. Two of the 9 reconnection events occurred at 23 minutes, after the first 20 minutes of observation. A protocol limited to 20 minutes would therefore have classified these patients as having persistent acute block. This finding is particularly relevant in light of the recent study by Patel et al., in which MI lesion transmurality was assessed using direct VoM epicardial electrograms and pacing over a 20- minute observation period (32). Their demonstration of time-dependent recovery of initially attenuated VoM electrograms provides a mechanistic framework for our observation that apparently complete bidirectional block may recover with continued observation. Our findings extend this concept by showing that conduction recovery may occur beyond 20 minutes even when VoM-EI is performed systematically before focal PFA. These observations support a standardized 30-minute waiting period when acute MI block stability is the procedural endpoint.

### Transmurality, Pseudoblock, and Acute Block Stability

An important caveat must frame the comparison with mechanistic studies (32). We did not cannulate the VoM for electrogram recording or pacing after ethanol infusion; therefore, bidirectional block in the present series was confirmed by conventional electrophysiological criteria and cannot be equated with proven transmural block (32). Patel et al. demonstrated the limitations of these criteria using direct VoM electrograms and pacing during MI PFA with FaraPulse, PulseSelect, and Affera (32). Durable attenuation of epicardial VoM electrograms after 20 minutes occurred in only 52.1% overall. Among the 31 Affera-treated patients, 29 initially showed VoM signal attenuation, but signals recovered in 8, leaving durable attenuation in 21 of 31 (67.7%); myocardial capture from the VoM persisted in 8 of 31 (25.8%); and pseudoblock was identified in 9 of 31 (29.0%). Apparent endocardial block after PFA therefore does not establish transmural interruption of the entire MI conduction network, and our 98.0% acute block rate should be interpreted as conventionally defined bidirectional block rather than proven transmural block.

With this caveat, comparison with the Affera subgroup of Patel et al. remains informative because the procedural sequences differed fundamentally. Patel et al. delivered PFA first and used VoM-EI when MI ablation remained incomplete, whereas we systematically performed VoM-EI before left atrial access and analyzed only patients in whom ethanol infusion was successful. In our cohort, block was obtained without CS ablation in 45 of 51 patients (88.2%), reflecting the combined effect of prior ethanol ablation of the Marshall substrate followed by focal endocardial PFA and not the efficacy of Sphere-9™ PFA alone. In the Affera subgroup of Patel et al., 18 of 31 patients achieved MI block with PFA alone; among the remaining patients subsequently treated with VoM-EI, 5 required additional CS ablation (32). Because the treatment sequence, definition of block, and use of direct epicardial interrogation differed, these proportions should not be interpreted as a comparative measure of efficacy.

Both studies nevertheless demonstrate time-dependent evolution of acute electrophysiological findings. In Patel et al., VoM electrograms initially attenuated after Affera PFA recovered in 8 of 29 patients (27.6%) during a 20-minute observation period without VoM-EI. In our study, conventionally defined bidirectional MI block recovered in 9 of 50 patients (18.0%) during 30- minute reassessment. These observations should not be considered quantitatively equivalent: Patel et al. measured recovery of epicardial VoM electrograms after PFA, whereas we measured recovery of MI conduction after systematic VoM-EI followed by PFA. Taken together, however, they emphasize that an apparently successful acute electrophysiological effect may evolve with time and that immediate assessment alone may overestimate lesion stability.

Our findings therefore complement rather than replicate those of Patel et al. Their study establishes that conventional block criteria may coexist with residual epicardial conduction and that PFA does not reliably produce transmural MI lesions. Our study addresses a different question: whether conventionally confirmed bidirectional block remains stable after systematic pretreatment of the Marshall substrate with ethanol followed by focal Sphere-9™ PFA. Despite this combined epicardial–endocardial strategy, conduction recovered in approximately one patient in five, and two thirds of these patients required additional CS ablation. Whether pretreatment of the Marshall substrate reduces early instability compared with a PFA-first strategy cannot be determined from these nonrandomized cohorts and would require direct VoM recordings and a prospective comparative design. What our data establish is narrower but clinically actionable: a conventionally accepted acute electrophysiological endpoint may be transient, and this recovery can be detected and corrected during the index procedure by protocol-defined reassessment.

### Acute Stability in the Context of Chronic PFA Line Durability

Invasive remapping studies have yielded markedly heterogeneous estimates of chronic MI-line durability. Reddy et al. reported durable MI block in 68% after a focal lattice-tip strategy with protocol-mandated remapping (28), whereas La Fazia et al. found persistent MI block in only 5.5% after pentaspline PFA with systematic 3-month remapping (29). Zaher et al. reported durable posterior MI block in only 2 of 9 patients undergoing clinically indicated repeat procedures after Sphere-9™ ablation, with epicardial gaps accounting for most reconnections (30). Brügger et al. observed anterior mitral line reconnection in 53% of remapped patients after PFA or RF (31). These estimates should not be interpreted as measurements of an intrinsic property of PFA because catheter configuration, waveform, line location, epicardial substrate, use of VoM-EI or CS ablation, and the timing and indication for remapping all influence observed durability.

Patel et al. provide complementary mechanistic evidence that endocardial MI lesions created with PFA, including Affera, are not uniformly transmural (32). Taken together, these observations emphasize that immediate block, acute block stability, and chronic lesion durability represent related but distinct electrophysiological endpoints. Our VoM-EI–first workflow intentionally modifies the Marshall epicardial substrate before focal PFA, and the standardized 30-minute waiting period interrogates the intermediate step between immediate procedural success and chronic durability. Our study does not establish chronic MI-line durability; rather, it identifies an early and potentially correctable component of conduction instability that would otherwise remain undetected at the index procedure. Whether waiting-period–guided re-ablation translates into improved chronic MI-line durability requires confirmation by protocol-mandated invasive remapping.

#### Comparison with Radiofrequency Based Strategies

In the randomized study by Zuo et al., acute MI reconnection after a 30-minute waiting period occurred in 36.1% with RF alone and 6.7% with RF plus VoM-EI (10). Our 18% rate lies between these figures and does not suggest that electroporation confers, per se, superior immediate lesion stability at the MI. Anatomical variability — including wall thickness, catheter–tissue orientation, CS musculature, and epicardial connections — may modify procedural difficulty, as previously demonstrated for cavotricuspid isthmus ablation (38). A randomized comparison of focal PFA and contemporary RF, both combined with standardized VoM-EI and block reassessment, is required.

### The Persistent Role of Coronary Sinus Conduction

Coronary sinus applications were delivered in 6 patients (11.8%) during the initial lesion set and in 6 patients after waiting-period conduction recovery, 3 of whom had already undergone CS ablation during the initial lesion set. Thus, 9 distinct patients (17.6%) underwent CS ablation during the procedure. Importantly, 6 of the 9 patients with early MI conduction recovery (66.7%) required targeted CS ablation to restore bidirectional block. Technically successful VoM-EI therefore does not exclude residual or recovered epicardial conduction outside the ethanol lesion footprint, and CS-related muscular connections may remain an important pathway for persistent or recovered MI conduction despite combined VoM-EI and endocardial PFA.

The potential importance of procedural sequence was raised before the PFA era. In the randomized study by Gillis et al., performing VoM-EI as the first step before RF ablation facilitated subsequent MI block and significantly reduced the endocardial RF burden compared with an RF-first strategy, although the sequence did not determine the final block rate after additional touch-up ablation (8). These findings provided a rationale for treating the Marshall epicardial substrate before endocardial energy delivery. Our study extends this concept to a focal PFA-based workflow but also shows that a VoM-EI–first strategy does not eliminate the potential contribution of CS-related conduction: two thirds of patients with waiting-period recovery ultimately required targeted CS ablation.

Patel et al. provide complementary mechanistic evidence in the PFA era. Residual CS-to-left atrial connections may persist despite PFA and VoM-EI and may require targeted CS ablation (32). In their Affera subgroup, 5 of 31 patients (16.1%) required CS ablation, compared with 6 of 51 patients (11.8%) during the initial lesion set in our cohort. This numerically lower requirement should not be interpreted as evidence of superiority of the VoM-EI–first sequence, given the indirect, nonrandomized comparison, different procedural sequences and electrophysiological endpoints, and restriction of our analysis to patients with successful VoM- EI. Differences in ethanol exposure may also have contributed: the median ethanol volume in our cohort was 9 mL (IQR 6–10; range 5–10), whereas Patel et al. reported a mean volume of 5.8 ± 3.2 mL in the Affera subgroup (32). Whether procedural sequence, ethanol exposure, MI anatomy, or other procedural factors influence the need for adjunctive CS ablation cannot be determined from these data.

Taken together, Gillis et al., Patel et al., and the present findings support a consistent procedural concept across thermal and PFA-based strategies: VoM-EI facilitates MI block and may reduce the subsequent endocardial ablation burden, but it does not eliminate the potential contribution of residual CS-related conduction. The CS should therefore be specifically interrogated when MI block cannot be achieved or when conduction recovers, while CS ablation should remain targeted rather than systematic. The particularly high requirement for CS ablation among patients with early recovery in our series (6/9, 66.7%) further suggests that waiting-period reconnection may identify a subgroup in whom residual or recovered CS-related conduction becomes electrophysiologically manifest only after apparently successful initial block.

#### Safety and Efficiency

No major complication occurred. The absence of coronary spasm despite focal PFA within the CS in a limited number of patients is reassuring but cannot establish safety, particularly because no systematic nitrate prophylaxis was used. A median skin-to-skin duration of 82 minutes for PVI, roof line, MI line, systematic VoM-EI, and a 30-minute waiting period indicates that protocol-defined reassessment remains compatible with an efficient workflow.

#### Clinical Implications

The practical endpoint of MI ablation should be stable rather than merely immediate bidirectional block. Patel et al. show why apparent block after PFA can be misleading by demonstrating incomplete transmurality and pseudoblock (32); our study shows how often conduction can recover despite a systematic VoM-EI–first strategy, how late that recovery can occur, and how it can be corrected during the index procedure. A structured sequence of VoM- EI, focal PFA, confirmation of bidirectional block, reassessment over a full 30-minute period, and targeted treatment of residual endocardial or CS conduction may therefore be more informative than judging efficacy from the energy source or immediate block alone.

#### Limitations

This study was single-center, single-arm, and observational, without an RF control group. The sample was modest and only 9 reconnection events occurred, precluding multivariable analysis. All procedures were performed by experienced operators at a high-volume center. The analysis was restricted to patients in whom VoM-EI was completed; the 4 patients in whom the vein could not be identified are therefore not represented, and the results describe the stability of MI block once the Marshall substrate has been treated rather than the performance of the strategy in an intention-to-treat population. Venographic non-identification of the vein does not exclude a muscular Marshall bundle along an obliterated vein, so no inference about epicardial substrate should be drawn from those 4 patients. We did not perform direct post-PFA VoM electrogram recording or VoM pacing as in Patel et al.; therefore, we cannot distinguish true transmural block from pseudoblock or assign individual reconnections to Marshall versus other epicardial pathways. Comparisons with Patel et al. are indirect and descriptive, between cohorts that differ in denominator, endpoint and interrogation window. Conduction recovery was assessed during a 30-minute interval, and no protocol-mandated chronic invasive remapping was performed. Consequently, acute stability cannot be equated with chronic durability or long-term freedom from perimitral flutter.

## CONCLUSIONS

Immediate bidirectional MI block was not synonymous with stable block. A VoM-EI–first strategy followed by Sphere-9™ focal PFA achieved acute block in 98% of patients, yet conduction recovered during the 30-minute waiting period in approximately one in five, two thirds of whom required targeted CS ablation, and recovery occurred as late as 23 minutes. These findings support a standardized waiting period with systematic reassessment and CS interrogation rather than reliance on immediate block. Systematic invasive remapping will be required to determine whether waiting-period–guided re-ablation translates into durable chronic block.

## DATA AVAILABILITY

The data supporting the findings of this study are available from the corresponding author upon reasonable request.

